# DeepPLL: Synchronization of non-invasive brain stimulation to deep brain stimulation

**DOI:** 10.64898/2026.06.17.26355884

**Authors:** Robert Toth, Nil Ramón i García, Mareike A. Gann, Monika Pötter-Nerger, Roger Dozen, Molly L. Zeitzschel, Andrew Sharott, Timothy Denison, Bettina C. Schwab

## Abstract

Deep brain stimulation (DBS) is increasingly viewed as a network-level intervention, yet clinical practice typically targets a single brain structure per hemisphere. Coordinated multi-site stimulation may help probe and modulate distributed circuits, but additional invasive implantation is limited by safety and ethical constraints. Here, we present an approach to couple DBS with non-invasive transcranial alternating current stimulation (tACS) via precise phase synchronization.

We introduce DeepPLL, an open-source interface device enabling real-time phase locking between DBS pulse trains and external stimulation like tACS. The system extracts DBS EEG artefacts using an isolated analogue front-end and stabilizes timing via a phase-locked loop (PLL) implemented in hardware or software. A digital phase-delay module with 1° resolution allows controlled adjustment of DBS phase, and low-jitter TTL outputs drive the external device.

In two individuals with Parkinson’s disease treated with subthalamic DBS, DeepPLL achieved reliable phase locking between DBS and motor-cortex tACS with sub-millisecond jitter in both PLL modes. This demonstrates feasibility of precise invasive-non-invasive stimulation synchronization in vivo and provides a platform for investigating phase-dependent network dynamics and plasticity.

---

Most invasive neuromodulation approaches target a single structure per hemisphere, such as the subthalamic nucleus (STN) or the internal globus pallidus (GPi) in Parkinson’s disease therapy. At the same time, deep brain stimulation (DBS) and other interventions are increasingly considered to operate at the network level, with widespread rearrangements of functional connectivity [1]. This raises the possibility that synchronized stimulation at multiple targets could help probe, or even shape desired network effects. However, exploratory implantation of electrodes at multiple targets is constrained by safety limits and ethical considerations.

Coupling non-invasive stimulation to DBS could therefore be an attractive alternative, allowing the exploration of additional stimulation sites without additional surgery or a potential increase in complications (Fig. 1A). In particular, transcranial alternating current stimulation (tACS) can be applied over many different cortical areas at a wide range of stimulation frequencies, including typical DBS frequencies (60–160 Hz) or frequencies of pathophysiological oscillations such as the beta band (13–30 Hz).

**Figure 1:**
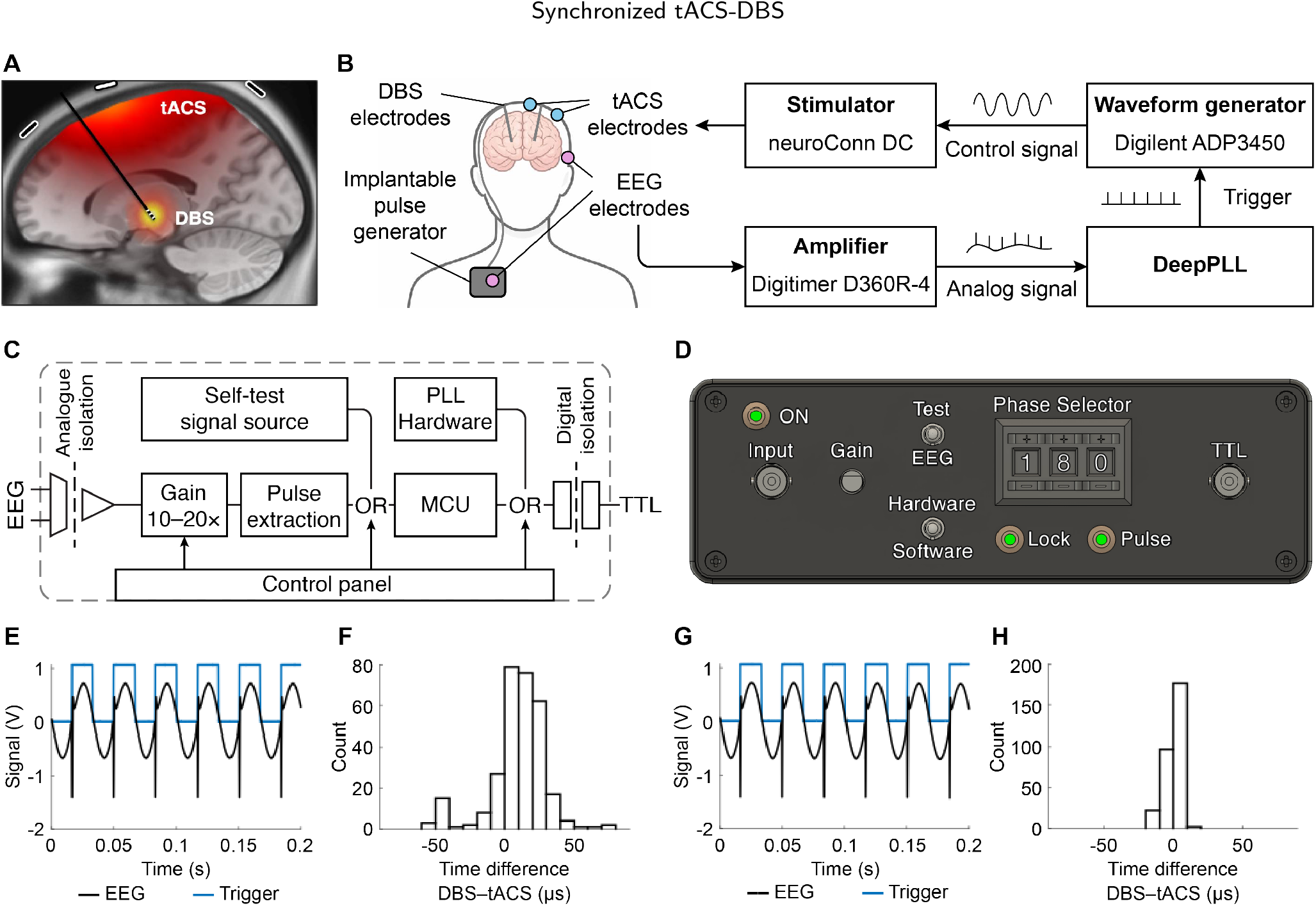
**A:** Conceptual illustration of coordinated tACS and DBS, targeting both cortical and sub-cortical areas. **B:** Experimental setup. DBS pulse artefacts were collected from an analogue EEG signal, and converted to stable digital trigger with selectable phase delays. The triggers drove a waveform generator and tACS stimulator. **C:** System overview. The DeepPLL contains an analogue signal conditioning circuit optimised to extract DBS pulse artefacts from an EEG background. The obtained pulse train is stabilised via a phase-locked loop (PLL). Users may select from a default hardware-based PLL, or a software-based PLL running in a real-time microcontroller unit (MCU). **D:** DeepPLL control panel. The EEG input and trigger ouput ports are isolated. Pre-amplifier gain is configurable. The device is equipped with a self-test mode through an internal pulse generator. The user may select between two PLL modes, and may enter the targeted phase delay. LED indicators mark successful phase-locking and output triggers. **E:** Example patient recording using the DeepPLL in hardware PLL mode. Black trace shows EEG containing artefacts of both DBS pulses and the tACS sine wave; blue trace shows the DeepPLL trigger output. **F:** Timing differences between DBS pulses and tACS triggers in hardware PLL mode. **G–H:** Corresponding recording and timing distribution for software PLL mode.

For example, synchronizing tACS over the motor cortex to DBS of the STN at a beta frequency would allow testing the pathophysiological relevance of this connection in Parkinson’s disease patients. If high-amplitude tACS and low-amplitude DBS each entrain a fraction of neurons in motor cortex and STN, respectively, then varying the phase lag between the two interventions should alter the relative timing of neuronal activity across these sites. Modulating functional connectivity at specific phase lags has the potential to differentially engage synaptic plasticity by changing the timing relationship between pre- and postsynaptic activity.

This principle of multi-site stimulation is already established in non-invasive stimulation, including dual-site tACS [2–5] and transcranial magnetic stimulation (TMS) [6, 7]. Combined TMS-DBS has also been shown to affect plasticity [8]. To our knowledge, however, phase-synchronised tACS-DBS has not been explored so far. Here, we present DeepPLL, an open-source interface device to synchronize DBS with external stimulation modalities (Fig. 1C–D). DeepPLL implements an isolated analogue front-end to reliably extract DBS pulse artefacts from surface electrode recordings; a selectable hardware or software phase-locked loop (PLL); and a digital phase delay selector with 1° resolution. Finally, the interface outputs isolated TTL triggers at the desired phase of the DBS cycle. The system is able to recognise DBS pulses as short as 5 µs at a 20 mV artefact magnitude.

While previous methods relied on fixed delay times to achieve multi-modal stimulation [8], our inclusion of a phase-locked loop ensures the combined stimulation pattern remains stable even in the presence of frequency drift in the DBS device. The DeepPLL system comes with two PLL options built-in: 1) a hardware PLL with a wide locking range of 2.5–150 Hz [9] and a lock-in time of 2.2 s; and 2) a software-based PLL (running in an embedded microcontroller) optimised for faster locking (0.75 s) over a narrower 15–120 Hz frequency range. The algorithm is open for user adjustments if other characteristics are experimentally desirable. Users may also disable the PLL function entirely to simply output the extracted DBS pulse times.

To validate the DeepPLL system *in vivo*, we conducted measurements in two patients with Parkinson’s disease who had bilateral DBS implants in the STN (Fig. 1B). For the duration of the experiment, the DBS devices were set to a frequency of 30 Hz, pulse width of 60 µs, and an amplitude of 1 mA on the left side, while the right side was disabled. Ipsilateral tACS was applied using a current-mode stimulator (neuroConn DC-Stimulator; neurocare group AG, Munich, Germany) with a 3-in-1 montage (central electrode: C3; outer electrodes: FC5, Cz, and CP5), a typical motor cortex montage. Stimulation intensity was set according to individual tolerance up to a limit of ±2 mA (4 mA peak-to-peak). Patient 1 reported uncomfortable sensations towards the higher end of the range, therefore the tACS amplitude was limited to ±0.8 mA. Patient 2 tolerated the full ±2 mA amplitude well. The participants were at rest during the sessions, no task was involved.

During stimulation, we monitored the EEG signal between channel P10 and an electrode in the infraclavicular region above the implanted pulse generator, using an analogue amplifier (D360R-4; Digitimer Ltd, Welwyn Garden City, UK), which served as the input to the DeepPLL system. Triggers from DeepPLL were sent to a waveform generator (ADP3450; Digilent Inc, Pullman, WA, US), which in turn provided the control signal to the tACS stimulator. The waveform generator was configured to deliver one period of a sinusoid pattern per trigger, allowing repeated triggering to present a nearly continuous tACS waveform. For validation, the analogue EEG signal was digitalized and saved at a rate of 200 kHz using an oscilloscope (ADP3450).

We achieved reliable phase locking of tACS to DBS in both patients, with both the hardware and software PLL modes. Fig. 1E–H shows the results for Patient 1, with jitter under 60 µs in hardware mode, and under 20 µs in software mode.

A current limitation of the system is that transcutaneous monitoring of DBS pulses requires monopolar stimulation, as bipolar artefacts may be too small for reliable detection [10]. Yet, monopolar stimulation remains the predominant DBS configuration. Furthermore, conventional DBS devices apply pulses to the left and right hemisphere at an interleaved timing, which could distort the phase locking when applied in typical clinical settings. Therefore, we restricted ourselves to unilateral DBS. Nevertheless, the software PLL leaves scope for future extensions, such as locking to every second DBS pulse, which could support use with bilateral DBS protocols.

In conclusion, we present a non-invasive solution to phase-lock tACS to DBS with high temporal precision. We show that this is even possible with low amplitudes of DBS (1 mA) and high amplitudes of tACS (up to ±2 mA). The design of our interface is made available to the research community as an open resource, and may serve as a starting point to investigate network dynamics and plasticity. Furthermore, our interface can also be used to synchronize sensory stimulation like visual, tactile or auditory input to a certain phase of DBS, allowing for combined sensory-electric network stimulation.

## CRediT authorship contribution statement

**Robert Toth:** Conceptualization, Methodology, Supervision, Writing – Original Draft; **Nil Ramón i García:** Formal analysis, Methodology, Software, Writing – Review & Editing; **Mareike A. Gann:** Investigation, Writing – Review & Editing; **Monika Pötter-Nerger:** Resources, Writing – Review & Editing; **Roger Dozen:** Investigation, Writing – Review & Editing; **Molly L. Zeitzschel:** Investigation, Writing – Review & Editing; **Andrew Sharott:** Conceptualization, Resources, Writing – Review & Editing; **Timothy Denison:** Conceptualization, Methodology, Writing – Review & Editing; **Bettina C**.

**Schwab:** Conceptualization, Funding acquisition, Methodology, Project administration, Resources, Supervision, Validation, Writing – Original Draft, Writing – Review & Editing.

## Ethical statement

The study was conducted in accordance with the Declaration of Helsinki. The study was registered in a public database before recruitment started (NCT07139093; https://clinicaltrials.gov). All participants provided their informed consent in writing. The study protocol was approved by the ethics committee of the Medical Association of Hamburg (PV7207).

## Data availability statement

The raw data and code required to reproduce the experimental findings of both patients are archived at https://doi.org/10.4121/12846417-2bb9-4f96-a025-09ec93604628. The design files required to recreate the DeepPLL system are available at https://github.com/roberttoth02/DeepPLL.

## References

[1] A. Horn, M. Reich, J. Vorwerk, N. Li, G. Wenzel, Q. Fang, …, and A. Kühn. Connectivity predicts deep brain stimulation outcome in parkinson disease. Annals of Neurology, 82(1):67–78, 2017.

[2] B. C. Schwab, J. Misselhorn, and A. K. Engel. Modulation of large-scale cortical coupling by transcranial alternating current stimulation. Brain Stimulation, 12(5):1187–1196, 2019.

[3] R. M. Reinhart and J. A. Nguyen. Working memory revived in older adults by synchronizing rhythmic brain circuits. Nature Neuroscience, 22(5):820–827, 2019.

[4] B. C. Schwab, P. König, and A. K. Engel. Spike-timing-dependent plasticity can account for connectivity aftereffects of dual-site transcranial alternating current stimulation. NeuroImage, 237:118179, 2021.

[5] M. A. Gann, I. Paparella, C. Zich, I. F. Grigoras, S. Huertas-Penen, S. W. Rieger, A. Thielscher, A. Sharott, C. J. Stagg, and B. C. Schwab. Dual-site beta transcranial alternating current stimulation during a bimanual coordination task modulates functional connectivity between motor areas. Brain stimulation, 18(5):1566–1578, 2025.

[6] L. P. Lafleur, S. Tremblay, K. Whittingstall, and J. F. Lepage. Assessment of effective connectivity and plasticity with dual-coil transcranial magnetic stimulation. Brain Stimulation, 9(3):347–355, 2016.

[7] S. Van Malderen, M. Hehl, S. Verstraelen, S. P. Swinnen, and K. Cuypers. Dual-site tms as a tool to probe effective interactions within the motor network: a review. Reviews in the Neurosciences, 34(2):129–221, 2023.

[8] K. Udupa, N. Bahl, Z. Ni, C. Gunraj, F. Mazzella, E. Moro, …, and R. Chen. Cortical plasticity induction by pairing subthalamic nucleus deep-brain stimulation and primary motor cortical transcranial magnetic stimulation in parkinson’s disease. Journal of Neuroscience, 36(2):396–404, 2016.

[9] R. Toth, A.B. Holt, M. Benjaber, A. Sharott, and T. Denison. Frequency and phase synchronization in distributed (implantable-transcutaneous)neural interfaces. 41st Annual International Conference of the IEEE Engineering in Medicine & Biology Society (EMBC), 2019.

[10] M. A. Iacono, S. R. Atefi, L. Mainardi, H. C. Walker, L. M. Angelone, and G. Bonmassar. A study on the feasibility of the deep brain stimulation (DBS) electrode localization based on scalp electric potential recordings. Frontiers in Physiology, 9(1788), 2019.

